# Comparing outcomes of ultra-low-cost hearing aids to programmable, refurbished hearing aids for adults with high frequency hearing loss in Malawi: A feasibility study

**DOI:** 10.1101/2023.03.08.23286971

**Authors:** Bhavisha Parmar, Mwanaisha Phiri, Louis Jailos, Regina Kachapila, Ben Seleb, Wakisa Mulwafu, Vinay Manchaiah, M. Saad Bhamla

## Abstract

**Introduction:** Access to ear and hearing health services are limited or non-existent in low-income countries, with less than 10% of the global production of hearing aids is distributed to this population. The aim of this feasibility study was to compare the outcomes of an ultra-low-cost hearing aid (LoCHAid) to programmable, refurbished hearing aids for adults with high-frequency hearing loss, in Blantyre, Malawi.

**Methods:** Sixteen adults with high frequency hearing loss, and no prior experience of hearing aids, took part in this study, nine were fitted with the LoCHAid and seven were fitted with refurbished, programmable hearing aids, for a one-month trial. Five standardized hearing qualities questionnaires were used to compare outcomes pre and post device fitting and between devices. Questionnaire scales were analysed using general linear models and inductive thematic analysis was used to evaluate qualitative data.

**Results:** Overall, there was no significant difference found between LoCHAid and refurbished hearing aids, and the two device types each showed a similar degree of improvement after fitting. Qualitative data identified two key themes: Sound Quality and User experience.

**Conclusion:** The results from this feasibility study are encouraging, but a comprehensive, larger clinical study is needed to draw firm conclusions about the LoCHAid’s performance. This study has identified key improvement indicators required to enhance sound quality and user experience of the LoCHAid.

## Introduction

Access to ear and hearing health services are limited or non-existent in low-income countries (Wilson et al., 2017), with less than 10% of the global production of hearing aids is distributed to this population (World Health Organisation, 2004). Age-Related Hearing Loss (ARHL) is one of the most prevalent chronic conditions in older adults and is estimated to affect 900 million people by 2050 (World Health Organisation, 2021). ARHL is managed primarily through use of hearing aids (Van Eyken et al., 2007), but uptake varies globally (Bainbridge & Ramachandran, 2014; Hartley et al., 2010).

One of the factors contributing to variable hearing aid uptake is cost. Hearing aids can cost between $500-$3000 in the US. The WHO guideline states that a hearing aid should be no more than 3% of the gross national product, per capita, per hearing aid (McPherson, 2011). Therefore, according to World Bank figures, for low-income, lower-middle income, and low- and middle-income countries, the affordable price would be approximately $20, $67.77, and $135, respectively. To address the growing need for hearing health, non-governmental organisations have been developing ear and hearing care services, and providing donated hearing aids, in low resource settings for many years (Newall et al., 2019; Thammaiah et al., 2017; Wertz et al., 2017). One of the key challenges in this process is the lack of trained local hearing healthcare professionals and a lack of specialised resources. Also, the large-scale fitting of donated hearing aids has several ethical implications. Engagement with local stakeholders is key for the sustainability of effective, patient centred hearing aid services (Kaspar et al., 2020).

### Ear and Hearing Care in Malawi

Malawi is a landlocked country located in southeastern Africa, sharing borders with Tanzania, Mozambique, and Zambia. It has a population of around 20 million people. Audiology and hearing healthcare services are extremely limited in Malawi (Mulwafu et al., 2018). There are only two publicly available audiology departments in the country, one in Blantyre and one in Lilongwe. Within the public hospital system, there are two ENT Specialist doctors; 32 ENT Clinical Officers; 4 audio technicians and one audiologist.

Hearing aids are not routinely provided by the Malawian Ministry of Health. Instead, the country relies on donated hearing aids provided by charitable organisations (Parmar et al., 2021). Despite donated hearing aids being available in cities including Lilongwe and Blantyre, much of Malawi’s ear and hearing care needs are underserved. Furthermore, refurbished hearing aids are pre-used devices, and this process relies on a constant flow of hearing aids and the relevant consumables and programming tools from the donation source, into Malawi. Another barrier to uptake is that digital hearing aids mainly function by using a battery which is not locally available. A retrospective study to understand the profile of patients attending the Queen Elizabeth Central Hospital Audiology department, Blantyre, Malawi found that demand for hearing healthcare services is growing in Malawi, but hearing aid uptake is low (Parmar et al., 2021). Of the 2,299 patients seen over a two-year period, 61% of adults were found to have some degree of hearing loss, but only 28% were fitted with refurbished hearing aids. Some patients had access to employment health insurance to pay towards the hearing aids, but others relied on self-funding. There is need for lower cost hearing device for Malawian ear and hearing care services to become more accessible and sustainable.

### The LoCHAid- an ultra low cost hearing aid for age related hearing loss

Preliminary data from both electroacoustic testing and simulated gain measurements demonstrates that a low-cost device, LoCHAid, provides amplification in the high-frequency that is needed for individuals mild- to moderate ARHL (Sinha et al., 2020). It is a minimal component hearing aid to address ARHL and has been adapted since the original study to improve the user experience and sound quality. The LoCHAid is a body worn, pre-set, rechargeable, hearing device. Headphones are used rather than ear moulds or tubes to couple to the patient’s ear. Due to the open-source nature of the device, it could be manufactured locally and could be offered to users with minimal cost. Although the original LoCHAid study confirmed the presence of the high frequency gain necessary to address ARHL, there is a need to clinically validate this technology, particularly in contexts where it may be of most use, i.e., low resource countries. Therefore, the primary aim of this feasibility study was to compare the outcomes of the LoCHAid to programmable, refurbished hearing aids in individuals with high-frequency hearing loss. In addition, we gathered user perspectives on these devices.

## Method

### Study Design and Ethical Considerations

This feasibility study involved the following phases: protocol development, participant recruitment, outcome measure translation, staff training and the clinical validation of the LoCHAid. The study was carried out at Queen Elizabeth Central Hospital Audiology department, Blantyre, Malawi.

Ethical approval was obtained from the Malawian College of Medicine Research and Ethics Committee (COMREC) (P.07/20/3091). Informed consent was obtained from all participants before the intervention was received. An information sheet was given to the participants, which outlined the purposes of the study. This was also summarized verbally before written consent was obtained.

### Recruitment

A purposive sampling strategy was implemented to select people over the age of 18 years with bilateral mild to moderate high frequency sensorineural hearing loss, and no experience of hearing aid use, and no reported cognitive or neurological conditions. Recruitment took place during outreach clinical activities or clinical activities within QECH Audiology. Audiology clinicians carried out diagnostic otoscopy, pure tone audiometry and tympanometry on all patients. All participants were reimbursed their travel costs and given a financial incentive to attend each research session.

### Staff training

The QECH audiology team were given remote and in-person training by a practicing audiologist (first author BP). Training included how to use the LoCHAid device, hearing aid fitting, hearing aid testing, counselling, study procedures, clinical testing, follow up and data management. Staff were in regular contact with the UK audiology team throughout the project. All QECH audiology technicians involved were also trained in basic research methods to aid the data collection process. All QECH staff assisted in creating the English and Chichewa hearing aid instruction manuals for the LoCHAid and the refurbished hearing aids.

### Outcome Measures

The five standardized hearing qualities or hearing aid benefit measures used in this project that were chosen after a joint discussion within the research team. The outcome measures were: Glasgow Hearing aid Benefit Questionnaire part one (GHABP) (Gatehouse, 1999), Speech, Spatial and Qualities of Hearing Scale (SSQ-12) (Noble et al., 2013), Listening effort Assessment Scale (EAS) (Alhanbali et al., 2017), Satisfaction with Amplification in Daily Life Questionnaire (SADL) (Cox & Alexander, 1999) International Outcome Inventory: Hearing Aids (IOI-HA) (Cox et al., 2000). The SADL is a fifteen-item questionnaire, divided into 4 subscales: positive effect, service and cost, negative features and personal image. However, an item referring to hearing aid cost and an item referred to telephone use were removed as the hearing aids were issued at no cost in this study, and the LoCHAid is not suitable to use with the telephone.

As all questionnaires were originally written and validated in English and the lack of hearing related outcome measures in the Malawian national language of Chichewa, the questionnaires were translated to Chichewa by a qualified Malawian translator with had previously translated for research studies (Manda-Taylor et al., 2022; Singal et al., 2021). Forward and backward translation processes were implemented (Hall et al., 2018), and the Malawian audiology team were involved to assisted to ensure accurate translation of audiology specific technical terms. When translating research questionnaires, it is important to assure congruency between words and their true meaning in the language to which the questionnaire is translated (Eremenco et al., 2005). The Chichewa questionnaires were trialed on a small number of normal hearing adults and necessary amendments were made to improve consistency, accuracy, and context. Open questions were also designed within a topic guide to explore participants’ experiences of their hearing devices. The topic guide included questions about the overall user experience, preference of usage for specific situations and barriers for continued usage.

### Hearing aids

Two types of hearing device were used in this feasibility study: a fixed-frequency-response low cost hearing aid (LoCHAid) (Sinha et al., 2020) and a refurbished, programmable hearing aid (Oticon). The LoCHAid has a fixed-frequency threshold, making it less tunable to individual participants’ hearing thresholds, but more suitable for health professionals who have no specialist ear and hearing care training to fit. The LoCHAid was first demonstrated by the audiology clinician before the participant listened through the device. The programmable hearing aids were donations from the charitable organisation Deaf Kidz International (DKI) and they were cleaned, checked and reset by audiology clinicians at a DKI partner organisation based in Lusaka, Zambia. All refurbished hearing aids used in this study were open fittings - coupled to the appropriate slim tubes and domes. They were programmed to participants’ hearing thresholds using the NAL NL1 prescription formula. Fine tuning adjustments were made in the consultation to ensure adequate audibility and comfort for each participant.

### Participants

Initially, 18 participants consented to take part in the study where nine were randomized to the LoCHAid group, and nine to the refurbished hearing aid group. After fitting 2 people from the refurbished hearing aid group withdrew from the study due to unforeseen circumstances. Therefore, sixteen participants with bilateral mild to moderate sensorineural hearing loss participated in this study. Demographics are described in the table 1. Hearing thresholds are presented in Figure 1.

**Table 1.** Participant demographics

| **Age range** | **Number of participants** |
| --- | --- |
| 18-30 | 1 |
| 31-50 | 3 |
| 51-60 | 1 |
| 60+ | 11 |
| **Sex** |  |
| Male | 11 |
| Female | 5 |
| **Work status** |  |
| Working | 4 |
| Not working | 12 |

**Figure 1.**
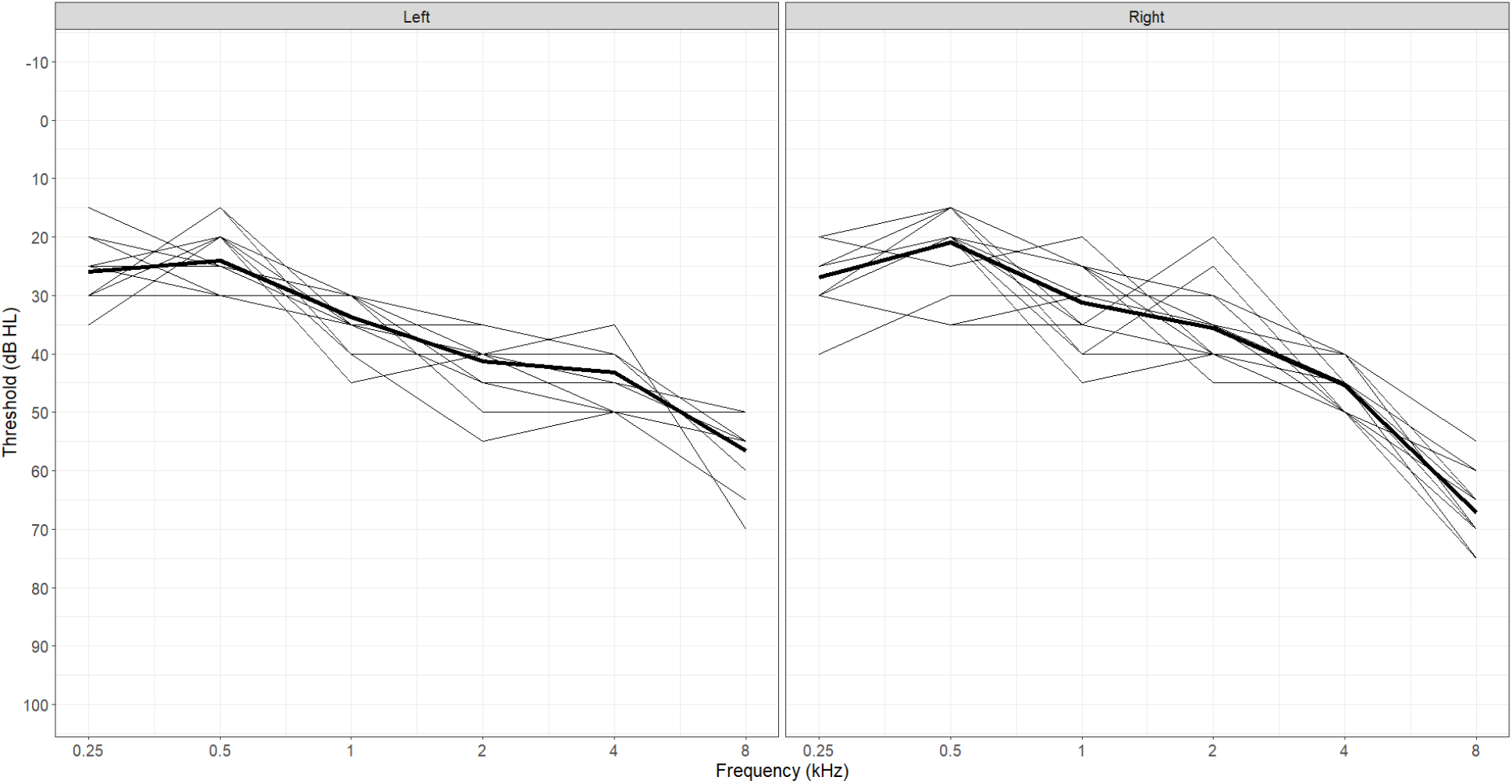
Hearing thresholds for all participants, presented for each ear. The bold line represents mean thresholds.

### Study protocol

Each participant completed three in-person visits to the QECH Audiology department for this study.

During visit 1 participants completed a diagnostic audiological assessment, including the following 4 questionnaires:

- Demographics Questionnaire
- Glasgow Hearing aid Benefit Questionnaire part one (GHABP) (Gatehouse, 1999)
- Speech, Spatial and Qualities of Hearing Scale (SSQ-12) (Noble et al., 2013)
- Listening effort Assessment Scale (EAS) (Alhanbali et al., 2017)

Participants were then randomly assigned to one of two groups: LoCHAid or refurbished hearing aids. The audiology clinicians carried out the hearing aid fittings using the allocated devices. Both groups were given detailed instruction booklets, specifically created for this study (written in Chichewa and English), to show them how to use the hearing aids and how to contact the audiology department. They were asked to use the hearing device as much as possible, in a variety of situations over the one-month trial. They were counseled on good communication tactics, realistic expectations, and acclimatization to the new sense of amplification, in line with typical clinical hearing aid fittings.

The follow up visit took place 4 weeks after the initial hearing aid fitting. The Chichewa versions of the following questionnaires were completed during the session:

- Glasgow hearing aid benefit questionnaire (GHABP) part 2
- Speech, Spatial and Qualities of Hearing Scale (SSQ-12)
- Listening effort Assessment Scale (EAS)
- Satisfaction with Amplification in Daily Life Questionnaire (SADL) (Cox & Alexander, 1999)
- International Outcome Inventory: Hearing Aids (IOI-HA) (Cox et al., 2000)

During the questionnaires, participants were asked to base their answers on their experience of hearing aid use within the one-month trial. After the follow up, participants were able to keep their trial hearing device if they wanted to. Additional open questions were asked in an interview style to explore the users’ general usage and experience of the allocated hearing devices.

A second follow up was carried out 3 months after fitting to check for medium term hearing aid use. During the session, open questions were asked, and notes were transcribed by the QECH team.

### Data Analysis

As recommended in the literature, the SSQ12 subscales (speech, spatial, and qualities) were calculated by averaging the scores of the four items in each category together. The GHABP disability subscale was calculated by averaging across the four disability scenarios. Improvement in hearing was evaluated by comparing the extent of initial disability (recorded upon inclusion before device implementation) and the extent of residual disability (recorded at follow-up after device implementation). The SADL was analysed by using individual items and its established subscales. The listening effort assessment scale and the IOI-HA were evaluated by global score differences and with individual item comparisons.

Statistical modeling was completed using R version 4.1.1. The SSQ12, GHABP, and Listening effort questionnaires were each completed both before and after hearing aid fitting by using linear mixed modelling for repeated measures. We tested each subscale as a dependent variable against changes in scores over time (before vs. after hearing aid fitting), between devices (LoCHAid vs. refurbished), and for an interaction between the two to indicate whether one device changed more than another over time. We additionally included a random grouping factor for participant to control for repeated measures. Repeated measures modeling was completed using the R package *lme4*. Satisfaction with amplification in daily life and IOI-HA items were each addressed individually, using a general linear model, with each item tested for differences between devices. Estimated marginal means were calculated using the R package *lsmeans (Lenth, 2016)*. For all models, residuals were confirmed as normally distributed using QQ-plots with the Kolmogorov-Shapiro test for normality. Multicollinearity among predictors was tested by calculating the variance inflation factor and was deemed negligible.

Inductive thematic analysis (Braun & Clarke, 2006) was carried out to analyse open text answers to questions including: “How would you describe your experience using the hearing device?”.

### Positionality

Hearing aid fitting and follow ups were conducted in Chichewa by train audio technicians (authors MP, LJ and RK). Their background as health professionals may have influenced the appointment dynamics with the participants. B.P (first author) was present in the clinic to assist the clinicians during the hearing device fitting appointments but did not make their presence known in case this influenced the flow of the appointments. B.P completed the data analysis in collaboration with the Malawian audiology team to ensure contextual details were not overlooked or misinterpreted.

## Results

Data from sixteen participants are presented in this study. Nine had been fitted with the LoCHAid device and seven wore bilateral refurbished hearing aids.

### Glasgow Hearing Aid Benefit Profile

Part one of the GHABP was completed at the first session, and part two was completed at the follow up session. Figure 2 displays the results of both parts, for both groups. Results were collapsed across the four questionnaire scenarios. The 6 dimensions of the GHABP: initial disability, initial handicap, HA use, HA benefits, residual disability, and HA satisfaction, for both devices can be seen in Figure 2. Overall, residual disability was reduced compared to the initial disability (subscale scores reduced from 2.56 (SE=0.18) to 1.53 (SE=0.18)). This was a significant reduction (F(1,13)=5.15, *p*=0.041). However, there was no significant difference found between devices (LoCHAid vs. Refurbished), and the two device types each showed a similar degree of improvement after fitting.

**Figure 2.**
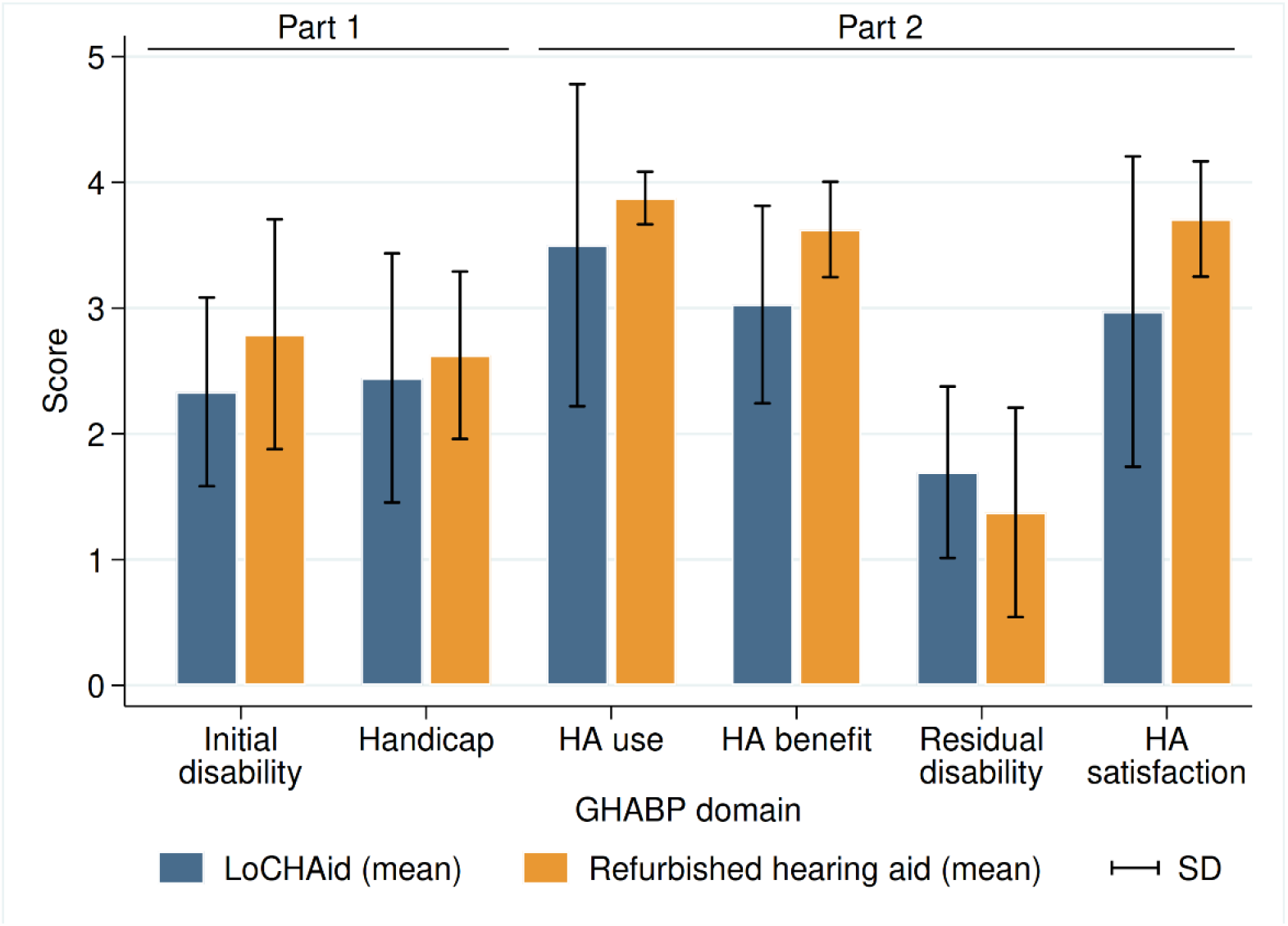
GHABP part one (pre fitting) and part two (post fitting) results. LoCHAid: n=9, refurbished hearing aid: n=7. Data collapsed across questions in each category with mean and standard deviation presented. Disability scores: 0 = no difficulty, 5= cannot manage at all.

### Speech, Spatial and Qualities of Hearing Scale (SSQ-12)

There was a significant improvement after device fitting in each of the speech (F(1,13)=13.24, *p*=0.003), spatial (F(1,13)=8.45, *p*=0.012) and qualities (F(1,13)=5.57, *p*=0.035) subscales, as shown in Figure 3. However, there was no significant difference between LoCHAid vs. refurbished devices, and the degree of improvement after fitting was similar when comparing the two.

**Figure 3.**
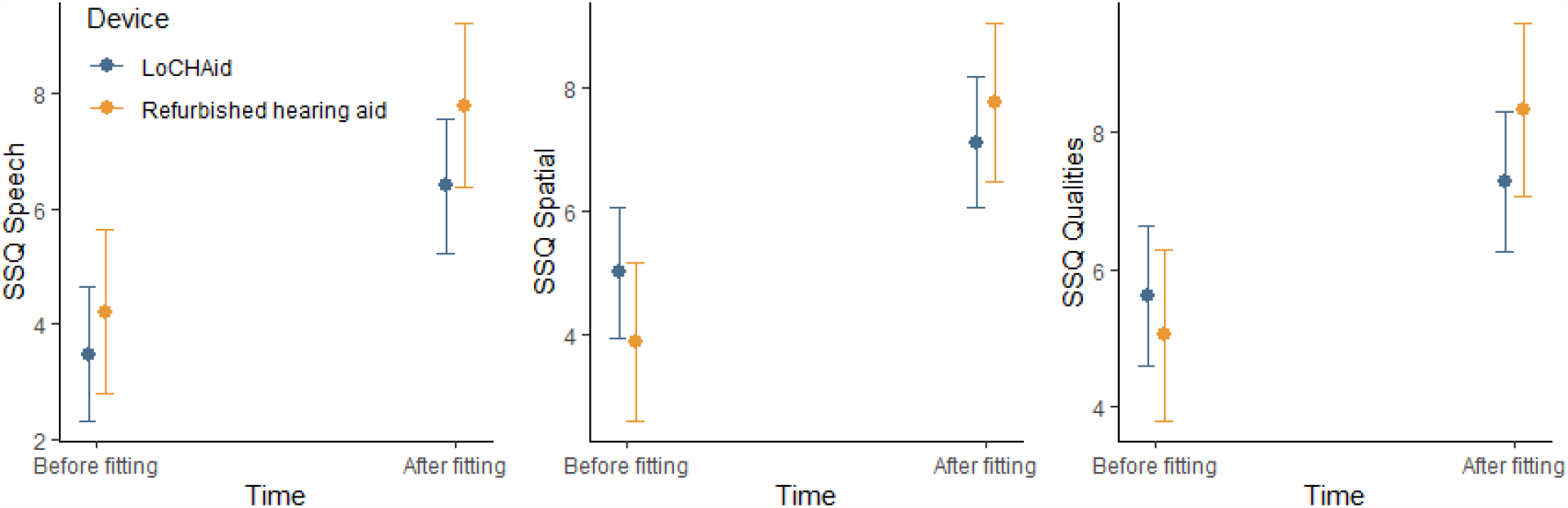
SSQ12 subscale scores before and after fitting for LoCHAid (n=9) and refurbished devices (n=7), with mean predicted scores and 95% confidence intervals shown. Category subscales 0= not at all, 10= perfectly.

### Listening Effort Assessment Scale (EAS)

Of the six items addressed in the listening effort questionnaire, five showed significant improvements over time, as outlined in Table 1. The only item which did not show a significant improvement was ‘*How easily can you ignore other sounds when trying to listen to something*’. Overall, there was no significant difference between LoCHAid vs. refurbished devices in any of the items, and the degree of improvement between time points was not significantly different between devices for any item. This same pattern was true for the global score, as shown in Table 2 and Figure 4.

**Table 2.** Listening effort questionnaire items before and after hearing aid fitting, with estimated scores and standard error shown for each item (high score indicates less effort). Significant differences are shown using asterisks (*, p<0.05).

| Item | Before | After | Significance |
| --- | --- | --- | --- |
| Listening effort in conversation | 4.64 (0.53) | 7.86 (0.53) | $F(1,13)=6.59, p=0.023 *$ |
| How much do you concentrate with listening to someone | 4.33 (0.55) | 7.75 (0.55) | $F(1,13)=11.39, p=0.005 *$ |
| How easily can you ignore other sounds when trying to listen to something | 4.92 (0.51) | 7.31 (0.51) | $F(1,13)=2.55, p=0.134$ |
| Do you have to put in a lot of effort to follow discussion in class/meeting/lecture | 5.06 (0.55) | 7.69 (0.55) | $F(1,13)=6.17, p=0.027 *$ |
| Do you have to put in a lot of effort to follow conversation in noise | 4.61 (0.50) | 7.50 (0.50) | $F(1,13)=5.61, p=0.034 *$ |
| Do you have to put in a lot of effort to listen on the telephone | 5.03 (0.55) | 8.06 (0.55) | $F(1,13)=7.37, p=0.018 *$ |
| Global score (combined average) | 4.76 (0.45) | 7.69 (0.45) | $F(1,13)=8.69, p=0.011 *$ |

**Figure 4.**
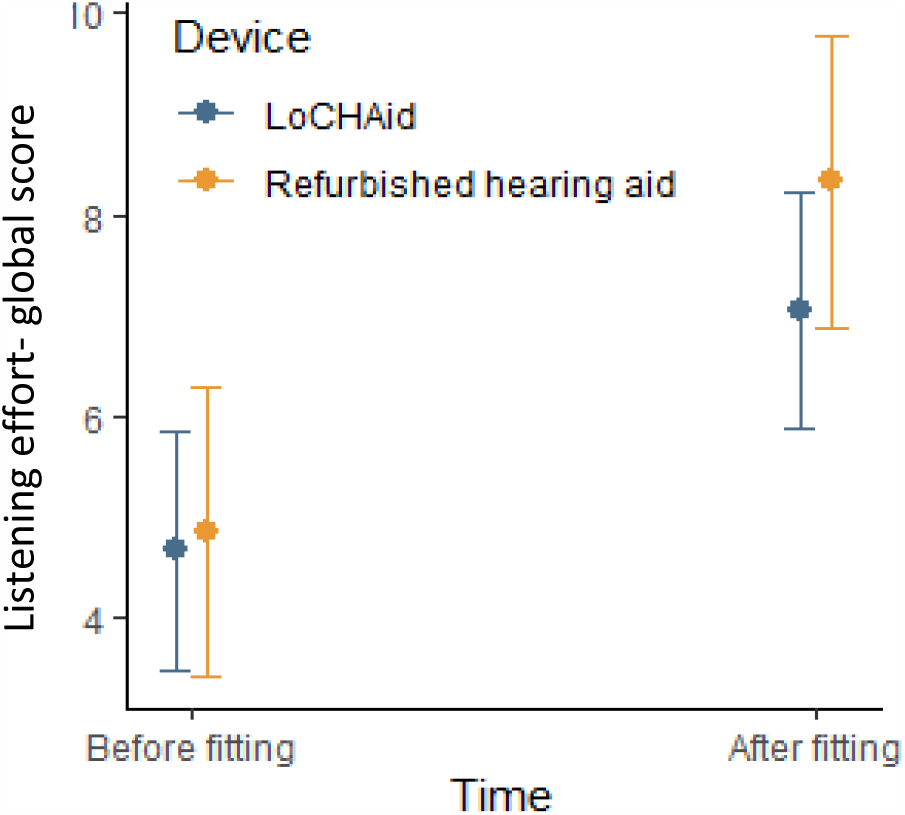
Listening effort questionnaire global score before and after fitting for LoCHAid and refurbished devices, with mean predicted scores and 95% confidence intervals shown.

### Satisfaction of amplification in daily life

Of the fifteen items in the Satisfaction with amplification in daily life questionnaire, four indicated a significant preference toward refurbished devices relative to LoCHAid devices. These items are shown in Table 3. Overall, participants found refurbished devices to be significantly more helpful with understanding people, more natural sounding, more dependable, and found their hearing aids to be more worth the trouble.

**Table 3.** Satisfaction with amplification questionnaire items for each device type, with estimated scores and standard error shown for each item. Significant differences are shown using asterisks (*, p<0.05).

|  | LoCHAid | Refurbished | Significance |
| --- | --- | --- | --- |
| *Compared to using no hearing aid at all, do your hearing aids help you understand people?* | 4.56 (0.50) | 6.33 (0.62) | t(13)=2.24, p=0.043 * |
| *Are you frustrated when your hearing aids pick up sounds that keep you from hearing what you want to hear?* | 2.44 (0.56) | 2.17 (0.69) | t(13)=-0.31, p=0.760 |
| *Are you convinced that obtaining your hearing aids was in your best interests?* | 5.33 (0.63) | 5.17 (0.77) | t(13)=-0.17, p=0.870 |
| *Do you think people notice your hearing loss more when you wear your hearing aids?* | 3.44 (0.62) | 4.83 (0.76) | t(13)=1.42, p=0.180 |
| *Do your hearing aids reduce the number of times you have to ask people to repeat?* | 4.78 (0.51) | 6.17 (0.62) | t(13)=1.72, p=0.108 |
| *Do you think your hearing aids are worth the trouble?* | 4.33 (0.43) | 6.33 (0.52) | t(13)=2.96, p=0.011 * |
| *Are you bothered by an inability to get enough loudness from your hearing aids without feedback?* | 2.56 (0.70) | 3.67 (0.86) | t(13)=1.00, p=0.335 |
| *How content are you with the appearance of your hearing aids?* | 5.00 (0.54) | 6.5 (0.66) | t(13)=1.77, p=0.100 |
| *Does wearing your hearing aids improve your self-confidence?* | 4.78 (0.56) | 6.17 (0.68) | t(13)=1.58, p=0.139 |
| *How natural is the sound from your hearing aids?* | 3.44 (0.39) | 6.00 (0.48) | t(13)=4.10, p=0.001 * |
| *How competent was the person who provided you with your hearing aids?* | 6.67 (0.20) | 6.83 (0.25) | t(13)=0.52, p=0.613 |
| *Do you think wearing your hearing aids makes you seem less capable?* | 3.67 (0.65) | 3.67 (0.80) | t(13)=0.01, p=1.000 |
| *How pleased are you with the dependability (how often they need repairs) of your hearing aids?* | 4.78 (0.27) | 6.17 (0.33) | t(13)=3.28, p=0.006 * |
| **Positive effect** | 4.54 (0.40) | 6.03 (0.49) | t(13)=11.34, p=0.035 * |
| **Service** | 5.72 (0.15) | 6.50 (0.18) | t(13)=3.33, p=0.005 * |
| **Negative features** | 2.50 (0.46) | 2.92 (0.56) | t(13)=0.573, p=0.576 |
| **Personal image** | 4.04 (0.38) | 5.00 (0.46) | t(13)=1.62, p=0.129 |

### International Outcome Inventory: Hearing Aids (IOI-HA)

Of the seven items in the IOI-HA questionnaire, only one showed a significant difference between device types. Users of refurbished devices reported significantly less difficulty than they used to have in the situations that they considered most important for requiring a hearing aid, compared to the LoCHAid users (see Table 4). The overall score comparison is shown in Figure 5.

**Table 4.** IOI-HA questionnaire items for each device type, with estimated scores and standard error shown for each item. Significant differences are shown using asterisks (*, p<0.05).

|  | LoCHAid | Refurbished | Significance |
| --- | --- | --- | --- |
| Think about how much you used your present hearing aid(s) over the past two weeks. On an average day, how many hours did you use the hearing aid(s)? | 3.78 (0.19) | 4.17 (0.24) | $t(13)=1.27, p=0.226$ |
| Think about the situation where you most wanted to hear better, before you got your present hearing aid(s). Over the past two weeks, how much has the hearing aid helped in that situation? | 3.67 (0.24) | 3.83 (0.30) | $t(13)=0.44, p=0.670$ |
| Think again about the situation where you most wanted to hear better. When you use your present hearing aid(s), how much difficulty do you STILL have in that situation? | 3.22 (0.25) | 2.0 (0.31) | $t(13)=-3.04, p=0.009 *$ |
| Considering everything, do you think your present hearing aid(s) is worth the trouble? | 3.44 (0.19) | 4.0 (0.23) | $t(13)=1.85, p=0.087$ |
| Over the past two weeks, with your present hearing aid(s), how much have your hearing difficulties affected the things you can do? | 2.67 (0.27) | 2.17 (0.34) | $t(13)=-1.15, p=0.271$ |
| Over the past two weeks, with your present hearing aid(s), how much do you think other people were bothered by your hearing difficulties? | 2.33 (0.30) | 2.17 (0.37) | $t(13)=-0.35, p=0.735$ |
| Considering everything, how much has your present hearing aid(s) changed your enjoyment of life? | 3.44 (0.21) | 3.83 (0.25) | $t(13)=1.18, p=0.258$ |
| Global score | 3.22 (0.09) | 3.17 (0.12) | $t(13)=0.37, p=0.716$ |

**Figure 5.**
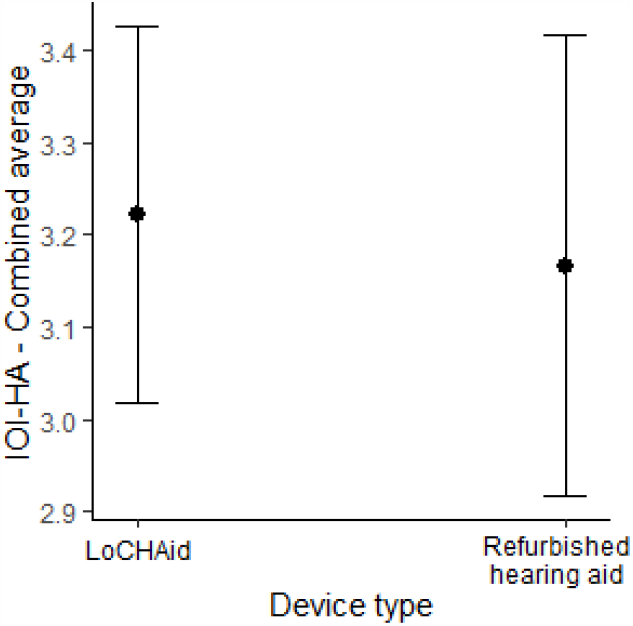
IOI-HA average scores for LoCHAid and refurbished devices with 95% confidence intervals shown.

### Qualitative data

Two key themes from the qualitative data obtained in the first follow up appointment are described below, with some example quotes. Table 5 summarises participant feedback from the second follow up session.

**Table 5.** Comments from second follow up appointment (3 months after hearing aid fitting)

| Participant | Device type | Still wearing hearing aids (Yes/No) | User experience |
| --- | --- | --- | --- |
| 1 | LoCHAid | No | The hearing aid make ears feel like they are blocked, and this makes it difficult to hear |
| 2 | Refurbished | Yes | The hearing aids are being helpful, and participant is wearing them regularly but often runs out of batteries. |
| 3 | Refurbished | Yes | The hearing aids are being helpful in most situations |
| 4 | LoCHAid | Yes | The hearing aid is being helpful but sometimes it makes ears itchy in his ears. The hearing aid also makes a humming noise which is disturbing in quiet places |
| 5 | LoCHAid | Yes | The hearing aid is being helpful but sometimes too noisy. Also, needs more volume when in challenging situations |
| 6 | LoCHAid | Yes | The hearing aid is working and is being helpful |
| 7 | LoCHAid | Yes | The hearing aid is working but to her the volume is very low and there is no volume adjustment option. |
| 8 | Refurbished | Yes | Positive experience using the hearing aids in a range of situations. Hearing aid is working |
| 9 | LoCHAid | Yes | The hearing aid is being helpful. Cannot wear the device in church or in meetings due to how the headphones look. |
| 10 | Refurbished | Yes | The hearing aid is working and is being helpful. |
| 11 | LoCHAid | Yes | With the hearing aid they can hear loud/medium level sounds but sound is not clear for soft sounds |
| 12 | LoCHAid | Yes | The hearing aid is working and is being helpful. |
| 13 | Refurbished | No | Sees no different between using hearing aids and without hearing aids. Stopped wearing the hearing aids after 2<sup>nd</sup> follow up. |
| 14 | Refurbished | Yes | The hearing aid is working and is being helpful. |
| 15 | Refurbished | not known | Did not attend follow up |
| 16 | LoCHAid | not known | Did not attend follow up |

#### Theme 1: Sound Quality

Participants commented on the sound quality of the amplification and the internal noise of the hearing device. Overall, all participants reported positive experiences of the refurbished hearing aid’s sound quality. However, those in the LoCHAid group reported that although the device improved their hearing ability, the internal noise from the device negatively impacted the listening experience.

*“Apart from amplifying sound, the sound it also produces is a noise like when you are tuning a radio”* (LoCHAid)

*“Volume control would be very helpful because sometimes I needed it higher”* (LoCHAid)

*“I liked everything about the sound”* (Refurbished)

*“I can hear much better with the device and I can hear the things I couldn’t hear without it” (*LoCHAid)

*“I think I hear perfectly but it took some time to get used to it”* (Refurbished)

*“too noisy”* (LoCHAid)

#### Theme 2: User Experience

Some participants liked the appearance of the LoCHAid as it did not look like a traditional hearing aid and therefore helped them avoid unnecessary attention. However, others felt the use of headphones made it look like they were using a music device, e.g., radio, and therefore not paying attention to their surroundings. The structure of the LoCHAid was reported to be quite delicate and there were some concerns raised about the number of visible open ports which may cause the device to malfunction in humid, dusty environment. Participants also commented on overall appearance and usability of each device.

*“The appearance is good, nobody knew it is hearing aid and that I have a hearing loss”* (LoCHAid)

*“It makes me look older”* (Refurbished)

“*Very easy to use*” (Refurbished)

*“Since it has headphones, you might wear it in places, you are not supposed to put headphones, people get disappointed in you.”* (LoCHAid)

*“The machine has lots of open spaces where dust can get in, like where the headphones go and the on button switch”* (LoCHAid)

*“Needs to be more powerful or have a volume control so I can control it, also sometimes headphones come out of my ear if I am eating/chewing”* (LoCHAid)

*“At church people thought I was listening to my phone or the radio. People thought because I am old that I am wearing this device to listen to music and look younger. Maybe they thought*

*I was being rude. It is a problem when it doesn’t look like something that is helping my medical condition”* (LoCHAid)

*“less visible headphones needed”* (LoCHAid)

“*The machine looks like it will break easily, what if I need a replacement or if the headphones stop working-can I use any headphones?”* (LoCHAid)

*“it is easy to put on and off, the colour of the headset makes it get dirty quickly. It improves my hearing, but the headphones can fall out easily. I don’t like being able to see inside the machine as I am worried it will break easily”* (LoCHAid)

## Discussion

This feasibility study is the first to clinically evaluate the effectiveness of the LoCHAid in a low resource setting. The results, from this Malawian population, found that the LoCHAid and the refurbished programmed hearing aids were similarly beneficial for people with high frequency hearing loss but that some improvements are required to improve the LoCHAid sound quality and user experience.

Low hearing aid uptake or hearing aid availability in Sub Saharan Africa has been noted in the literature (Hlayisi & Ramma, 2019; Parmar et al., 2021). Furthermore, many countries rely on hearing aid donations from non-governmental organisations (Seelman & Werner, 2014; Thiga et al., 2022). A number of previous studies have explored the effectiveness of low cost hearing aids (Parving & Christensen, 2004) and some research has piloted their use in low resource settings (Vo et al., 2018), as recommended in the WHO Guidelines for hearing aids and services for developing countries (World Health Organisation, 2004). Pienaar et al (2010) found positive patient benefit of hearing aid fitting in a South African context, even without optimal hearing aid fittings (suboptimal fitting due to financial constraints) (Pienaar et al., 2010). McPherson and Wong (2005) used the IOI-HA to investigate the effectiveness of an affordable (approximately $125USD) pre-programmed hearing aid in Hong Kong, with most patients benefitting from the hearing aids. Qualitative interviews in the study found the main disadvantages of the device to be hearing aid design (e.g. difficult to change battery) and hearing aid related problems including feedback and internal noise. Borg et al (2017) compared low-cost hearing aid fitting in the community to a health centre approach in Bangladesh. The trial demonstrated similar hearing aid benefit for both approaches in five out of seven of the IOI-HA items (Borg et al., 2018). However, the low-cost hearing aids used in many of these studies are still unaffordable for many low resource countries.

The present study found the LoCHAid had potential to serve people with hearing loss in Malawi and broadly performed similarly to digitally programmed refurbished, donated behind-the-ear hearing aids. Three standardized measures, SSQ-12, GHABP and EAS, were carried out before and after hearing aid fitting to explore the effectiveness of hearing aid use on various listening situations. Results from each of these outcome measures found that both devices, the LoCHAid and refurbished hearing aid, improved hearing abilities and listening effort (compared to the unaided experiences) at a similar degree. Two outcome measures reviewed overall hearing aid use and experience and were completed at the follow up appointment. Results from the SADL found that the LoCHAid was less helpful compared to the refurbished hearing aid in helping to understand people during conversation, the LoCHAid was less natural sounding and less dependable overall. Results from the IOI-HA found that users of refurbished devices reported significantly less difficulty, compared to LoCHAid, than they used to have in the situations that they considered most important for requiring a hearing aid.

During the follow up visits, participants were asked open questions about their overall hearing aid use and experience and probing questions about how their device does or does not benefit them. This feedback was analysed to present key themes and summarized to identify some key improvement indicators required to improve the LoCHAid user experience. These improvement indicators included factors affecting the design, hearing aid output, features and accessories and are shown in Table 6. If these improvement indicators are actioned, the user experience would be improved, and this is likely to increase device use and device reliability.

**Table 6.** Improvement indicators for LoCHAid

| Area of evaluation | Summary of feedback | Actionable changes |
| --- | --- | --- |
| Design | Dislike viewing device circuitry and easily builds up dust | Replace transparent panel |
|  | • Use of headphones causes others to think the user is not attentive to conversation (as they may be listening to music/radio)<br>• Gaps in device e.g headphone port could let dust in | • Close any unnecessary ports to ensure device is more resistant to dust/humidity<br>• Use of less visible headphones |
| Output | • Internal noise too high<br>• After charging, volume seemed higher | • Trial different headphones to achieve optimal output<br>• Reduce internal noise |
| Features | • Low output and volume control need | • Explore whether volume control could be added without compromising output/consistency |
| Accessories | • Headphone port seem loose, and functionality depends on how much the cables are pushed in | • More port needed |

The study had some limitations. Firstly, due to the nature of the device and its role in providing only high frequency amplification, finding the appropriate patient population for this trial was challenging. Despite running many outreach recruitment events, in neighboring villages and towns, the number with mild-moderate high frequency hearing loss, with no other otological symptoms was low. This project took place during the COVID-19 pandemic and was affected by many delays due to travel restrictions, procurement delays and staff shortages in Malawi. Our team were able to overcome these challenges by carrying out regular remote training sessions between our team in the UK and the Malawian audiology team. Also, we arranged for an audio technician from another city in Malawi to work at QECH for the duration of this feasibility trial. A key strength of this study was the involvement of the Malawian audiology clinicians as they were trained to recruit and consent participants, run the feasibility trial and had key involvement in the development of the outcome measures and the hearing aid instruction booklets. This experience and training would help future trials that take place in the same centre.

There are currently no validated hearing aid outcome measures (speech perception tests or questionnaires) in Malawi. However, during the development phase of this study five hearing qualities/hearing aid benefit questionnaires were translated to Chichewa and validated with a small normal hearing adult population. Backwards and forwards translation was implemented by a professional English-Chichewa translation service and the audiology team at QECH cross checked all translation. Additional validation of these Chichewa questionnaires using a larger population would further confirm the consistency of the measures. Speech perception and speech in noise perception measures are used to guide and evaluate hearing aid fittings in audiology settings (Parmar et al., 2022), but Chichewa measures of this kind are not currently available. A Digits in Noise test, or similar, could be used in future for this purpose, if self-reported English-competence and age were considered (Potgieter et al., 2018).

## Conclusion and future research

The results from this feasibility study are encouraging, but a comprehensive, larger clinical study is needed to draw firm conclusions about the LoCHAid’s performance. This study has identified key improvement indicators required to enhance sound quality and user experience of the LoCHAid. Once these improvements are made, further electroacoustic, speech perception and self-report testing should be completed on similar patient populations.

## Data Availability

All data produced in the present study are available upon reasonable request to the authors

## Notes

### Competing Interest Statement

The authors have declared no competing interest.

### Funding Statement

This study was funded by the Global Development Incubator

